# Morning Report Improves Residents’ Diagnostic Competence & Clinical Problem-Solving Ability

**DOI:** 10.1101/2022.11.02.22281868

**Authors:** Muhammad Tariq, Maham Vaqar, Shahab Abid, Wasim Jafri

## Abstract

**Introduction:** Morning report (MR) is an educational activity that uses inpatient case-based teaching. Given the rapid changes taking place in medical practice, it is important to assess the residents’ perspective regarding this teaching method.

**Objective:** To establish the perspective of residents in internal medicine on various aspects of MR and propose a format based on our observations.

**Study Design:** Observational cross-sectional study.

**Place & Duration of Study:** Data was collected from groups of residents in the Department of Medicine at the Aga Khan University Hospital, from July 2002 to August 2007.

**Methodology:** An observational cross-sectional survey on MR was conducted among the residents of the Department of Medicine at Aga Khan University. A 22-item questionnaire was distributed among the residents based on the purpose, format, and contents of the morning report, as well as the most appropriate person to present and conduct it, and how frequently they should be carried out.

Analyses were carried out using the statistical software ‘Statistical Package for Social Sciences’ (SPSS)

**Results:** 92% of residents believed MR to be an effective teaching activity with 65% of them choosing ‘Improvement in clinical problem-solving ability’ as the primary purpose of MR followed by ‘improving presentation skills’ (62%) and ‘conveying medical knowledge to the residents’ (58%). 79 residents (87%) believed that the junior resident should present the case history. 75 residents (83%) thought that faculty on call at time of patient’s admission should conduct MR. Residents wanted to discuss diagnostic work-up (90%) and management (89%) of specific interesting cases (79%) in MR.

**Conclusion:** MR is an effective educational activity and should be an essential component of any post-graduate residency program within the country and outside.

## Introduction

It is vital to establish the Morning Report as an important teaching component for residency programs throughout the world. It can potentially be an effective forum for both teaching and learning [1]. Morning Report is an activity that involves the presentation of cases of patients that are admitted through the Emergency Room (ER) and later discussed by the attending and post-graduate trainees. It functions as an instructive case-based teaching conference capable of providing a broad coverage of topics. [4, 5] The cases discussed are variable, and thus a great number of topics can be discussed in a setting that caters to all post-graduate trainees simultaneously.

The Morning Report can serve many purposes, including but not limited to conveying medical knowledge, reviewing management decisions, evaluating residents’ performance, helping the chief of service to keep track of developments, and, most importantly, case-oriented teaching. The Morning Report has been described as the intellectual highlight of the day [6]. It has several educational benefits for the residents as it is an interactive teaching session and involves learners across multiple levels, with an emphasis on collaborative case discussion and active learning [8].

Since this mode of imparting medical education has not been well established, there has been discrepancy regarding the ideal format and goals of this activity. We here aim to assess the views of residents regarding the morning report in order to come up with suggestions for a more conducive and effective morning report practice.

Morning Report at the Aga Khan University Hospital, Karachi, Pakistan is a 60-minute conference held twice every week, in which cases of two to three patients admitted the previous night through the ER are presented. All postgraduate trainees on ward rotations are required to attend. The post-call faculty conducts the report, and usually an intern or a junior resident presents these cases, supported by the senior resident of the team.

Diagnostic dilemmas, difficult management, rare illnesses, and unusual presentations of common diseases are preferred for presentation. It is a stepwise presentation of history, physical examination, and laboratory workup followed by a problem-oriented interactive discussion on the differential diagnosis and management.

In an earlier study [9] conducted at our institution, the Morning Report was confirmed to be an effective teaching activity, and there was significant concordance in the opinions of faculty and residents regarding the aforementioned features of the morning report.

## Objective

To make our learning more learner-based and model it according to the needs and expectations of our residents, we sought to establish the perspective of residents on various aspects of MR and to propose a format based on our observations.

## Methodology

An Observational Cross-Sectional Survey was conducted in the Department of Medicine of the Aga Khan University Hospital, Karachi, Pakistan. Data was collected from groups of residents, from July 2002 to August 2007. A 22-item questionnaire was distributed among the post-graduate trainees after the Morning Report session and the participants are asked to return the printed questionnaire in four weeks. Written informed consent was obtained from all the trainees who completed the survey. Individual responses were kept confidential. The questionnaire was based on a 0–5 Likert scale and was divided into multiple stems seeking opinion on the purpose, format, content, frequency, and appropriate presenters of the morning report. The responses were categorized as follows; 0 to 3 as negative responses, and 4 and 5 as positive responses.

Means and standard deviations were calculated for continuous variables and frequencies for categorical variables. Univariate analysis was performed using the Pearson Chi-square test. Analyses were carried out using the statistical software ‘Statistical Package for Social Sciences’ (SPSS).

The study was exempted from Ethical Review Committee approval by the Institutional Review Board at the Aga Khan University and the data obtained were analyzed anonymously.

## Results

Out of the one hundred and fifteen post-graduate trainees who were given the questionnaire, 89 responded. Their mean age was 27.29 years (95% CI; 26.70-27.89) with 56% males and 44% females. Of the participants, 26% were interns, 24% were level-I residents, 17% were level-II residents, 12% were level-III residents, 9% were level-IV residents, and 12% were working as resident medical officers.

‘Improvement in clinical problem-solving ability’ was rated as the primary purpose of the morning report by 65% of the residents, followed by ‘improvement in presentation skills’ (62%) and ‘conveying medical knowledge to the residents’ (58%). ‘Evaluating resident’s performance’ (19%) and ‘inspiring clinical research’ (12%) were not ranked highly.

The idea of the post-call faculty conducting the morning report was favored by 83% of residents. A general internist was preferred by 57% and similar response rate was observed for the Chief Resident (56%) conducting the report. However, medical sub-specialists (36%) and the Chairman of the Department of Medicine (23%) were not preferred to conduct the report. (Table 1)

**Table 1:** Who should conduct and present morning report?

| Parameters | Results (positive response) |  |  |  |
| --- | --- | --- | --- | --- |
|  | Overall<br>89 Residents<br>N (%) | 2002<br>Residents<br>N (%) | 2007<br>Residents<br>N (%) | p-value |
| Who should Conduct Morning Report? |  |  |  |  |
| Chief Resident | 50 (56) | 26 (54) | 24 (59) | 0.679 |
| Post call faculty | 75 (83) | 44 (92) | 31 (76) | 0.038 |
| Chairman Dept. of<br>Medicine | 21 (23) | 15 (31) | 6 (15) | 0.066 |
| Internist | 51 (57) | 29 (60) | 22 (54) | 0.521 |
| Medical sub-specialist | 32 (36) | 21 (44) | 11 (27) | 0.097 |
| Fellow | 36 (40) | 18 (38) | 18 (44) | 0.540 |
| Who should Present Morning Report? |  |  |  |  |
| Junior Resident | 78 (87) | 42 (88) | 36 (88) | 0.965 |
| Senior Resident | 38 (42) | 19 (40) | 18 (46) | 0.521 |
| Intern | 66 (73) | 38 (79) | 28 (68) | 0.243 |
| Medical Student | 12 (13) | 5 (10) | 7 (17) | 0.359 |
| Faculty | 10 (11) | 7 (15) | 3 (7) | 0.279 |

Most of the residents favored discussion of specific interesting cases (79%), which would be helpful in their post-graduate examinations (69%). ‘Bed-side teaching’ was rated low by the participants (51%) as the format of morning report. A similar response was observed for the ‘distribution of handouts’ (43%). (Table 2)

**Table 2:** Format and contents of morning report.

| Parameters | Results (positive response) |  |  | p-value |
| --- | --- | --- | --- | --- |
|  | Overall | 2002 | 2007 |  |
|  | 89 Residents<br>N (%) | Residents<br>N (%) | Residents<br>N (%) |  |
| Format of Morning Report |  |  |  |  |
| Review specific interesting cases | 71 (79) | 35 (73) | 36 (88) | 0.081 |
| Review only last night’s admissions | 30 (33) | 19 (40) | 11 (27) | 0.205 |
| Review admission since last report | 28 (31) | 13 (27) | 15 (37) | 0.336 |
| Presentation on a set format | 28 (31) | 16 (33) | 12 (29) | 0.681 |
| Free presentation with time limit | 25 (28) | 12 (25) | 13 (32) | 0.483 |
| Distribute Journal articles | 35 (39) | 22 (46) | 13 (32) | 0.174 |
| Review for post-grad examination | 62 (69) | 35 (73) | 27 (66) | 0.470 |
| Distribute handouts | 39 (43) | 26 (54) | 13 (32) | 0.033 |
| Bedside teaching | 46 (51) | 31 (65) | 15 (37) | 0.008 |
| Contents of Morning Report |  |  |  |  |
| Diagnostic workup | 81 (90) | 45 (94) | 36 (88) | 0.328 |
| Disease process | 63 (70) | 35 (73) | 28 (68) | 0.633 |
| Tests and procedures | 61 (68) | 34 (71) | 27 (66) | 0.614 |
| Evidence based medicine | 62 (69) | 35 (73) | 27 (66) | 0.470 |
| Screening and prevention | 43 (47) | 24 (50) | 19 (46) | 0.731 |
| Medical Ethics | 37 (41) | 22 (46) | 15 (37) | 0.378 |
| Research methods | 29 (32) | 16 (33) | 13 (32) | 0.870 |
| Management issues | 80 (89) | 43 (90) | 37 (90) | 0.918 |

Diagnostic workup (90%) and management issues (89%) were rated highly by the residents as important elements to be covered in the morning report. Disease processes (70%) and evidence-based medicine (69%) were rated similarly. The residents showed low positive responses for discussions of medical ethics, screening and prevention, and research methods. (Table 2)

Most of the residents (87%) thought that the junior resident should present the patient history, while 73% of the residents favored the intern. All participants rated medical students and faculty very low as the presenter. (Table 1)

A large majority (89%) thought that all residents and interns should attend the activity. There were relatively low response rates regarding medical students, fellows, and faculty.

The majority were in favor of morning report occurring twice a week (68%) followed by weekly (17%) and daily (14%). The preferred timings were 8 to 9 am in the morning (82%) and 13 to 14 pm in the afternoon (10%).

Overall, it was considered to be an effective teaching activity by 92% of the participants.

On comparing the data collected in 2002 and in 2007, we found that residents’ opinions regarding Morning Report did not change significantly with time (Table 1 & 2). The few significant differences between the groups were improvement in clinical problem-solving ability (p=0.030), post-call faculty conducting the morning report (p=0.038), distribution of handouts (p=0.033) and bedside teaching (p=0.008) during morning report. (Table 1, 2)

## Discussion

Morning Report can become a staple for internal medicine residency programs for the years to come as it involves a diverse group of teachers and trainees with different learning objectives [2, 10, 11]. It is perceived as an instructive conference [12], where information is exchanged [4] while emphasizing inpatient medical learning[10, 13].

During this survey, we examined the perspectives of the residents on various aspects of the morning report. Our results showed that ‘improvement in clinical problem-solving ability’ and ‘presentation skills’ were highlighted by the residents as the main purpose of the morning report, followed by ‘conveying medical knowledge to the residents’. This was outlined in the previous studies on the morning report, which perceived medical education as its primary purpose [1, 4, 13].

Morning Report is not intended to inspire clinical research, as it is a case-based clinical teaching activity. This is reflected by the negative preferences of participants in our survey. Similarly, the idea of using a morning report to evaluate residents’ performance [10] was not highly favored by the participants. Residents wanted post-call faculty, preferably a general internist, to conduct the morning report. This is consistent with earlier studies [11].

Also, a majority of the responders believed that discussing management issues with diagnostic workup of specific interesting cases should be the norm for the morning report, whereas previous studies have suggested that the selection and mode of presentation of cases varies greatly among programs and tends to reflect the chief resident’s and attending physician’s preferences [14]. Also, participants favored morning meetings over bedside sessions, an observation similar to what Stickrath et al. reported in their study where attending rounds were found to be more useful than bed-side learning.[15]

In addition, though in our research, the majority of the responders believed that handouts were not useful, Luciano et al. in his study reported that a toolkit describing expectations, outlining teaching plans, and containing feedback forms improves this activity. [16]

At our institution, a stepwise presentation of the complete history, physical examination, and laboratory findings is followed by a problem-oriented discussion on differential diagnosis and management issues. It has been postulated that this approach not only makes the discussion more interesting but also fosters clinical problem-solving skills [5, 17,18].

A majority of the participants expressed the wish that the discussions should be directed towards their post-graduate examination, while most of the studies done earlier did not emphasize this need.

The residents favored a junior resident or an intern as the presenter of the patients at morning report [18] and they all agreed that morning report should be attended by all the residents, medical officers, interns, post-call faculty, and the chief resident. This would allow residents and the faculty to interact in an academic environment where residents may encounter potential role models [19-21].

Overall, our participants considered the Morning Report to be an effective teaching activity, as outlined by an earlier study [22], and felt that it was worthwhile to spend an hour, twice weekly, on this activity. This activity should be dynamic and, if conducted with a positive attitude and sufficient interpersonal rapport like other teaching activities, can produce excellent results [23]. Recently, the new idea of a “morning report blog” was found to be of real benefit if used in conjunction with case-based learning sessions [24].

## Conclusion

Morning Report is an effective teaching activity which improves the clinical problem-solving ability, presentation skills, and knowledge base of the postgraduate trainees. Post-call faculty, preferably a general internist, should direct the report, as opposed to a medical sub-specialist. It is an activity where diagnostic workup and management of specific interesting cases are learned. However, it is not a place for learning medical ethics, research methods or evaluating residents’ performance.

## Data Availability

The data underlying the results presented in the study are available from the Aga Khan University on reasonable request.

## Acknowledgements

The authors would like to acknowledge the contributions of Dr. Roger Sutton for his extensive review of the article and Safia Awan (Research Coordinator, Department of Medicine, Aga Khan University) for analyzing data in this study.

The authors also declare that this study was not supported or funded by any grant from the university or any other source.

